# Single Dose Vaccination in Healthcare Workers Previously Infected with SARS-CoV-2

**DOI:** 10.1101/2021.01.30.21250843

**Authors:** Saman Saadat, Zahra Rikhtegaran Tehrani, James Logue, Michelle Newman, Matthew B. Frieman, Anthony D. Harris, Mohammad M. Sajadi

**Author notes:** Co-senior authors (Anthony D. Harris, Mohammad M. Sajadi).

## Abstract

Coronavirus disease 2019 (COVID-19) vaccine shortages have led some experts and countries to consider untested dosing regimens. We studied antibody responses to a single dose of the Pfizer-BioNTech or Moderna vaccines in healthcare workers (HCW) with laboratory-confirmed COVID-19 infection and compared to them to antibody responses of HCW who were IgG negative to SARS-CoV-2 spike protein. HCW with prior COVID-19 showed clear secondary antibody responses to vaccination with IgG spike binding titers rapidly increasing by 7 days and peaking by days 10 and 14 post-vaccination. At all time points tested, HCW with prior COVID-19 infection showed statistically significant higher antibody titers of binding and functional antibody compared to HCW without prior COVID-19 infection (p<.0001for each of the time points tested). In times of vaccine shortage, and until correlates of protection are identified, our findings preliminarily suggest the following strategy as more evidence-based: a) a single dose of vaccine for patients already having had laboratory-confirmed COVID-19; and b) patients who have had laboratory-confirmed COVID-19 can be placed lower on the vaccination priority list.

---

Although vaccination is a safe and effective method of prevention of coronavirus disease 2019 (COVID-19) [1], current shortages in vaccine production and distribution have led some experts and countries to consider untested regimens (one or half dose of a vaccine). Persons infected with COVID-19 are thought to have protective immunity and memory responses [2] for at least 6 months; however, neither recall responses nor ideal dosing has been studied in those previously infected with COVID-19. We compared antibody responses to COVID-19 vaccination in healthcare workers (HCW) with and without previous COVID-19 infection.

## Methods

HCW who had previously enrolled in a hospital-wide serosurvey study [3], conducted from July to August, 2020 at the University of Maryland Medical Center were randomly contacted based on stratification into three groups: Group 1: IgG antibody negative; Group 2: IgG positive and with asymptomatic COVID-19; and Group 3: IgG positive with history of symptomatic COVID-19. Participants were vaccinated with either the Pfizer-BioNTech or Moderna vaccine, depending on personal preference and availability. Blood was drawn at Day 0 (or baseline), 7, 10, and 14 post-vaccination in December 2020, and January 2021 (draws could be +/- 1 day from assigned day). Plasma was tested for IgG ELISA to spike trimer, which was modified from an assay we have previously published [4] to give a read-out of half-maximal binding titers, as well as end-point binding titers. Day 0 and 14 samples of vaccinees were also tested for SARS-CoV-2 neutralization by live virus neutralization, as previously published [5]. As controls, samples from inpatients and outpatients infected with COVID-19 were included, chosen at 1 and 2 months after onset of COVID-19 symptoms, when antibody titers would be at their peak. Statistical analysis was carried out with GraphPad Prism 5 (GraphPad Software, San Diego, CA). Antibody titers between groups were tested by the 2-tailed Mann-Whitney test, with a p<.05 being considered significant.

## Results

A total of 3816 HCW were enrolled in the serosurvey study [3], of which 151 were randomly contacted, and 59 volunteers enrolled: 17 in the Group 1, 16 in the Group 2, and 26 in Group 3 (demographic details in Table 1). The median age was 38 for the Group 1, 40 for the Group 2, and 38 for Group 3. The percent females were 71% for Group 1, 75% for Group 2, and 88% for Group 3. For the IgG ELISAs, at each of the Days 0, 7, 10, and 14 for ELISAs, COVID-antibody positive HCW (Group 2 and 3) had higher half-maximal and end-point IgG titers than Group 1 (p<.0001). At day 14 the reciprocal half-maximal binding titers were 924 in Group 1, 34,033 in Group 2, and 35,460 Group 3. At Days 0 and 14, each of Groups 2 and 3 had higher ID99 live virus neutralization titers than Group 1 (p <.0001 for each). At 14 days, the reciprocal ID99 neutralization titers were 80 for Group 1, 40,960 for Group 2, and 40,960 in Group 3. Figure 1 provides a summary of the above findings.

**Table 1.** Study population baseline characteristics.

|  |  | **Group 1**<br>**(n=17)** | **Group 2**<br>**(n=16)** | **Group 3**<br>**(n=26)** | **Inpatient**<br>**(n=19)** | **Outpatient**<br>**(n=19)** |
| --- | --- | --- | --- | --- | --- | --- |
| Age (years) | Median | 38 | 40 | 38 | 41 | 32 |
|  | Range | 29-55 | 25-72 | 23-59 | 28-48 | 18 - 81 |
| Gender [n (%)] | Male | 5 (29) | 4 (25) | 3 (12) | 16 (84) | 8 (42) |
|  | Female | 12 (71) | 12 (75) | 23 (88) | 3 (16) | 11 (58) |
| Race/Ethnicity [n (%)] | Black or African American | 2 (12) | 5 (31) | 5 (19) | 4 (22) | 2 (11) |
|  | White or Caucasian | 12 (71) | 9 (56) | 18 (69) | 1 (6) | 13 (68) |
|  | Asian | 3 (18) | 2 (13) | 3 (14) | 0 (0) | 2 (11) |
|  | Hispanic or Latino | 0 (0) | 0 (0) | 0 (0) | 13 (72) | 2 (11) |
| Vaccine [n (%)] | Pfizer | 10 (59) | 6 (37) | 13 (50) | N/A | N/A |
|  | Moderna | 7 (41) | 10 (63) | 13 (50) | N/A | N/A |
| Days symptomatic | Median | N/A | 0 | 10 | N/A | N/A |
|  | Range | N/A | 0-2 | 3-34 | N/A | N/A |
| Hospitalization [n (%)] |  | N/A | 0 (0) | 4 (15) | 19 (100) | 0 (0) |
| Days from symptom onset | Median | N/A | N/A | N/A | 37 | 48 |
|  | Range | N/A | N/A | N/A | 28-55 | 29-63 |
| Months from Covid PCR+<br>to vaccination | Median (percent PCR tested) | N/A | 9.0 (44%) | 8.0 (73%) | N/A | N/A |
|  | Range | N/A | 6.5-9.7 | 6.0-10.3 | N/A | N/A |
| Months from Covid IgG+<br>to vaccination | Median | N/A | 6.2 | 6.1 | N/A | N/A |
|  | Range | N/A | 4.8-7.1 | 3.8-7.2 | N/A | N/A |

N/A-not applicable

**Figure 1.**
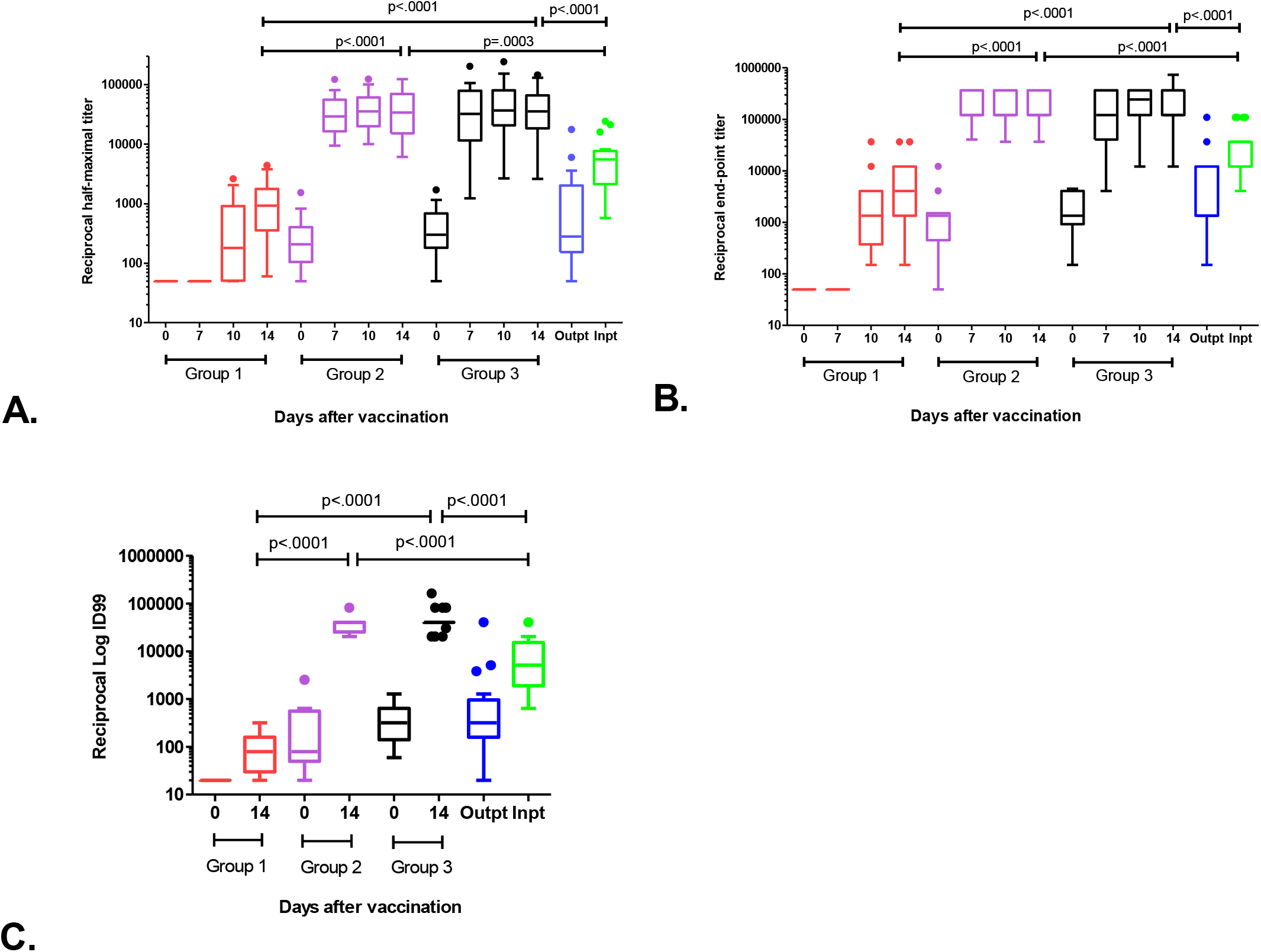
Anti-SARS-CoV-2 antibody responses after single dose of vaccination. After COVID-19 vaccination, plasma was drawn at 0, 7, 10, and 14 days and IgG binding titers against spike measured by ELISA and live virus neutralization at Day 0 and 14 measured. Single time point was also measured among COVID-19 outpatients or inpatients whose blood was drawn during peak antibody production (1-2 months after onset of symptoms). **A)** IgG spike trimer half-maximal titers. By 7 days and continuing to 14 days post vaccination HCW with prior COVID-19 who received a single vaccination developed higher peak IgG titers than HCW without COVID and COVID-19 infected patients. Half-maximal binding titers represent the dilution of plasma that achieves 50% of maximal binding of a known control that reaches saturation. **B)** IgG spike trimer end-point titers. By 7 days and continuing to 14 days post vaccination HCW with prior COVID-19 who received a single vaccination developed higher peak IgG titers than HCW without COVID and COVID-19 infected patients. End-point binding titers represent the highest dilution of plasma that that gives a reading above the statistically defined cutoff. **C)** Live virus neutralization. Serial dilutions of plasma was incubated for one hour with 100TCID50 of WA-1 strain SARS-CoV-2. This admixture was added to Vero cells, and cytopathic effect was assayed using live/dead staining at 72 hours. At 14 days, HCW with prior COVID-19 who received a single vaccination developed higher neutralization titers than HCW without COVID and COVID-19 infected patients. Small box size and single line seen in Groups 2 and 3 due to large numbers of values at 40,960. ID99 defined as highest dilution at which 99% cells were protected. Group 1= SARS-CoV-2 IgG negative healthcare worker (HCW). Group 2= asymptomatic SARS-CoV-2 IgG positive HCW. Group 3= symptomatic SARS-CoV-2 IgG positive HCW. Box plots represent 25% to 75% percentile, with individual dots representing outliers using Tukey’s method (1.5 x IQR).

## Discussion

We found that HCW previously infected with SARS-CoV-2, diagnosed via IgG to spike protein, had a classic secondary response to a single inoculation with a spike-based mRNA vaccine. That is, antibody titers started peaking at 7 days, and achieved higher titers and neutralization in 14 days compared to volunteers exposed to SARS-CoV-2 spike protein for the first time. Thus, there was an increase in both binding and functional antibodies (neutralization titers were 512 times larger). This occurred for HCW who were both symptomatic and asymptomatic with their SARS-COV-2 infection. Although we did not have peak titers for these individuals after natural infection, the titers developed after single vaccination was higher than peak titers in inpatients and outpatients with COVID-19, similar to what has been described in primary vaccination after 2 doses of the spike-based mRNA vaccines [6]. This secondary response occurs through activation of memory B cells [2]; and this study demonstrates recall responses through the antibody production stages in response to vaccination.

Given the above findings that one dose of vaccine can elicit a recall response, what we currently know about the duration of protective and memory responses, and until correlated of protection are identified, we think that in times of vaccine shortage, our findings preliminarily suggest the following strategy as more evidence-based: a) a single dose of vaccine for patients already having had laboratory-confirmed COVID-19; b) patients who have had laboratory-confirmed COVID-19 can be placed lower on the vaccination priority list; and c) lengthening time from Covid-19 infection to vaccination beyond the currently recommended 3 months.

## Data Availability

Data is available upon request.

## Acknowledgements

We would like to thank all of the study participants who donated their time and samples. ADH supported by CDC grant U01CK000556-02-01.

